# Whole genome sequencing improves tissue of origin diagnosis and treatment options for cancer of unknown primary

**DOI:** 10.1101/2024.08.09.24311642

**Authors:** Richard J. Rebello, Atara Posner, Ruining Dong, Owen W.J. Prall, Tharani Sivakumaran, Camilla B. Mitchell, Aidan Flynn, Alex Caneborg, Catherine Mitchell, Sehrish Kanwal, Clare Fedele, Samantha Webb, Krista Fisher, Hui-Li Wong, Shiva Balachander, Wenying Zhu, Shannon Nicolson, Voula Dimitriadis, Nicholas Wilcken, Anna DeFazio, Bo Gao, Madhu Singh, Ian Collins, Christopher Steer, Mark Warren, Narayan Karanth, Huiling Xu, Andrew Fellowes, Rodney J. Hicks, Kym Pham Stewart, Charles Shale, Peter Priestley, Sarah-Jane Dawson, Joseph H.A. Vissers, Stephen B. Fox, Penelope Schofield, David Bowtell, Oliver Hofmann, Sean M. Grimmond, Linda Mileshkin, Richard W. Tothill

## Abstract

Genomics holds promise for precision treatment and identifying the primary tissue of origin (TOO) in cases of cancer of unknown primary (CUP). We evaluated the feasibility and diagnostic superiority of whole genome and transcriptome sequencing (WGTS) over conventional panel testing in 72 patients using routine pathology samples. WGTS not only detected all reportable mutations identified by panel testing but also uncovered additional clinically relevant features in 76% of cases. Utilizing a CUP prediction algorithm (CUPPA) trained on WGTS data of known primary cancers, WGTS informed TOO in 77% of cases. Importantly, WGTS suggested potential treatment options for 79% of patients, a significant improvement over the 62% informed by panel testing. Additionally, WGTS and CUPPA applied to 22 cell-free DNA samples yielded high-likelihood TOO predictions in 41% of cases. These findings demonstrate that WGTS is diagnostically superior to panel testing, broadens treatment options, and is feasible using archived tissues and cell-free DNA.

## Main

Cancers of unknown primary (CUP) are metastatic tumours for which a tissue of origin (TOO) cannot be identified after standardized diagnostic investigations^1^. Although improvements in cancer diagnostics has likely reduced the incidence of CUP in recent years, it still accounts for 1-3% of all new cancer diagnoses^2^. While a minority (∼20%) of CUP patients have a favourable outcome corresponding to treatment responsive cancer types, most CUP patients have an unfavourable prognosis and empirical chemotherapy is less effective^1^.

DNA sequencing can identify therapeutically actionable mutations in a third or more of CUP tumours^3^. The CUPISCO clinical trial showed that patients with unfavourable CUP who received first-line molecular or immune-checkpoint targeting therapies based on tumour profiling data, had improved progression-free survival following three cycles of induction chemotherapy compared to platinum-chemotherapy treated controls^4^. Therefore, applying molecularly targeted treatments in CUP patients irrespective of the cancer origin is plausible, but despite this, targeted treatments can still show variable efficacy between cancer types^5^, while drug access through standard of care or clinical trials, is still often restricted to specific cancer types. Genomic tests that can resolve both the primary cancer diagnosis and direct targeted treatments are therefore needed.

Genomic profiling has been used to predict primary TOO in CUP, with a number of studies utilizing RNA expression^6–8^, DNA methylation profiling^9^ or DNA mutation profiling^10–12^. The application of DNA mutation data for TOO diagnosis is attractive as the same data also informs precision treatments. Some gene-specific mutations are enriched among certain cancer types, while the detection of mutational signatures, such as those associated with ultraviolet light and tobacco smoking, can also provide additional diagnostic evidence^13^. Indeed, the detection of diagnostic features using panel sequencing was shown to assist in resolving a likely TOO in a third of CUP patients^14^. Applying whole-genome sequencing (WGS) is expected to have superior diagnostic yield among CUP as the number of genome-wide features captured is much greater. One particularly promising application of WGS is the use of machine learning to predict TOO by using a combination of genome-wide driver and passenger mutation features, many of which cannot be reliably detected using panel sequencing^10,11,15^. Despite this, no systematic comparison of WGS to panel testing has been described specifically in CUP to date, and in particular when applying WGS to often degraded clinical material obtained from formalin-fixed paraffin-embedded (FFPE) samples^11,16^.

Here, we describe the feasibility of combining whole-genome and transcriptome sequencing (WGTS) in a series of 73 CUP tumours for 72 patients recruited to an Australian CUP clinical genomics study. For 59/73 (81%) tumours, we utilised FFPE samples and for 71/73 (97%) we also performed comprehensive cancer panel testing, enabling a direct comparison of reportable features between platforms. A CUP Prediction Algorithm (CUPPA)^11^ was applied to the WGTS data to compare the algorithmic TOO predictions against a pathologist’s favoured diagnosis who was informed by individual curated genome features. Finally, given tissue availability can be limiting factor for molecular profiling in many CUP patients we evaluated WGS of plasma cell-free DNA from CUP patients where high levels of circulating tumour DNA (ctDNA) were detected in a patient’s blood. We show the potential to use the WGS on cell-free DNA and the CUPPA method to confidently classify primary TOO when no tissue was available.

## Results

### Feasibility of clinical WGTS applied to a national CUP program

To assess the feasibility of WGTS for CUP patients we originally recruited 74 patients from a national CUP genomics study, where 61/74 (82%) cases had an unfavourable CUP prognosis according to EMSO criteria (Supplementary data 1). One case (CUP1209) had two clonally independent metastases based on discordant genomic profiles, indicating two synchronous and unrelated tumours, therefore a total of 75 tumours were sequenced. However, sequencing data was ultimately only useable for 73/75 tumours (discussed below). DNA cancer panel testing was done prior to attempting WGTS for 73/75 (97%) tumours using either a custom 386-gene cancer panel (CCP^14^), which involved sequencing both tumour and matched germline DNA (27/73, 37%), or a 523-gene tumour-only cancer panel (Illumina TSO500) (46/73, 63%), which also involved RNA-seq capture for fusion detection (Fig. 1a).

**Figure 1.**
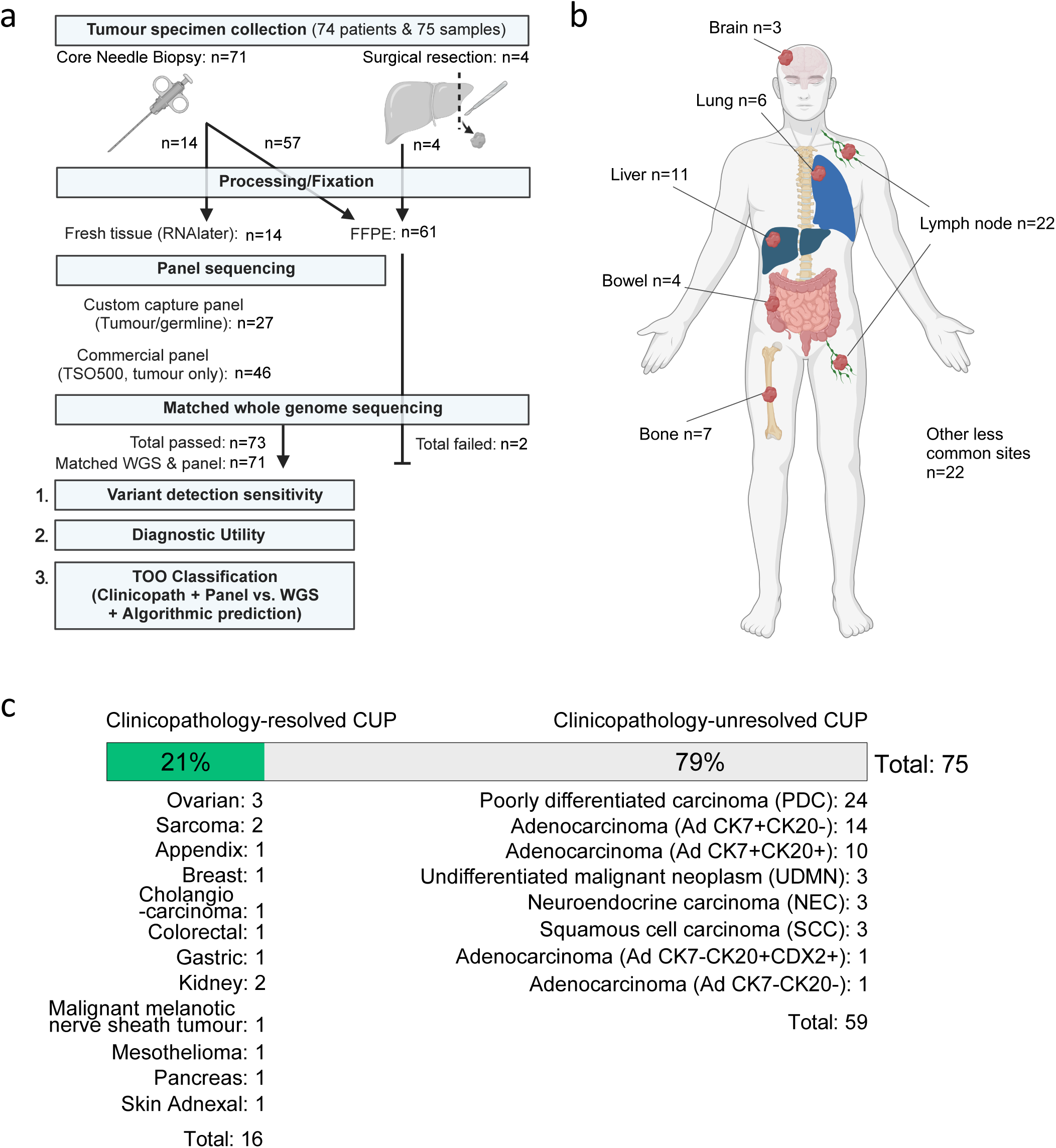
Selection of cancer of unknown primary cases for whole genome and transcriptome sequencing. **(a)** Flow chart of CUP cases profiled in this study. Tumour samples were either diagnostic core needle biopsies or surgical resections accessed from pathology archives as FFPE or fresh. Seventy-five samples from 74 CUP patients had WGTS, and 73 tumours had matched panel: 27 custom and 46 commercial cancer panels. **(b)** Patient anatomical sites sampled for molecular profiling in 75 CUP patients. Non-recurrent sites were combined as “other less common sites” (n=22). **(c)** Fraction of CUP tumours resolved (clinicopathology-resolved) or unresolved (clinicopathology-unresolved) pre-genomic testing, after a centralised pathology review with unresolved CUP categorized based on a modified OncoTree classification.

Most tumours (61/75, 81%) were FFPE tissues (Fig. 1a, Supplementary data 2); however, fresh tissue was sequenced for 14/75 (19%) cases and in three cases we had both fresh and FFPE material available for comparison. Most tissues were core needle biopsies (71/75, 95%), with the most common biopsy sites being lymph node or liver, representing 22/75 (29%) and 11/75 (15%) of cases respectively (Fig. 1b, Supplementary data 1).

Retrospective review of clinical and pathology information was performed by a single pathologist (OWJP) who initially favoured a likely single site diagnosis in 16/75 (21%) tumours before the genomic data was made available and these cases were termed clinicopathology-resolved (Fig. 1c). The remainder were clinicopathology-unresolved and were assigned a modified Memorial Sloane Kettering (MSK) OncoTree classification using CK7, CK20 and CDX2 immunohistochemistry to define CUP subsets as previously described^14^. The most common histological subtypes were adenocarcinoma (Ad) and poorly differentiated carcinoma (PDC), representing 26/75 (35%) and 24/75 (32%) of tumours, respectively (Fig. 1c). Neuroendocrine carcinoma (NEC, 3/75, 4%) and squamous cell carcinoma of unknown origin (SCC, 3/75, 4%) were minor subsets. (Fig. 1c, Supplementary data 1).

### Benchmarking FFPE to fresh tissue WGTS data

FFPE tissues are known to pose technical challenges for WGTS because of DNA damage sustained during fixation thus reducing DNA quality^17^. To minimize potential WGTS failure due to poor-quality FFPE DNA, we used a modified version of a PCR-based assay^18^ to determine the proportion of amplifiable DNA fragment sizes, excluding any cases that fell below a specified threshold (see methods). Only two tumour samples that passed initial PCR-based DNA quality control had unusable, poor-quality sequencing data and these were therefore excluded, leaving a total of 73 tumour samples from 72 patients for analysis. (Fig. 1a-c). Whole transcriptome RNA-sequencing was also unsuccessful for four tumours, but this did not preclude WGS DNA analysis and curation.

As expected, we observed that FFPE-derived DNA sequencing libraries had shorter median DNA fragment lengths than those derived from fresh tissues (Fig. 2a, FFPE: median of 437 base pairs (bp), range of 384 bp vs. Fresh: median of 618 bp, range of 245 bp) as well as a lower library complexity, indicated by higher sequence duplication rates (Supplementary data 2, FFPE: median of 25%, range of 56% vs Fresh: median of 7%, range of 15%). On average, 35% more sequencing data was generated for FFPE samples (Fig. 2b, median: 2.5×10^9^ reads, range 1.5-4×10^9^ reads) compared with fresh tissue samples (median: 2×10^9^ reads, range: 1.7-2.5×10^9^ reads), although the target median sequence coverage (60x sequencing depth) was still not reached for three FFPE tumours (Fig. 2c).

**Figure 2.**
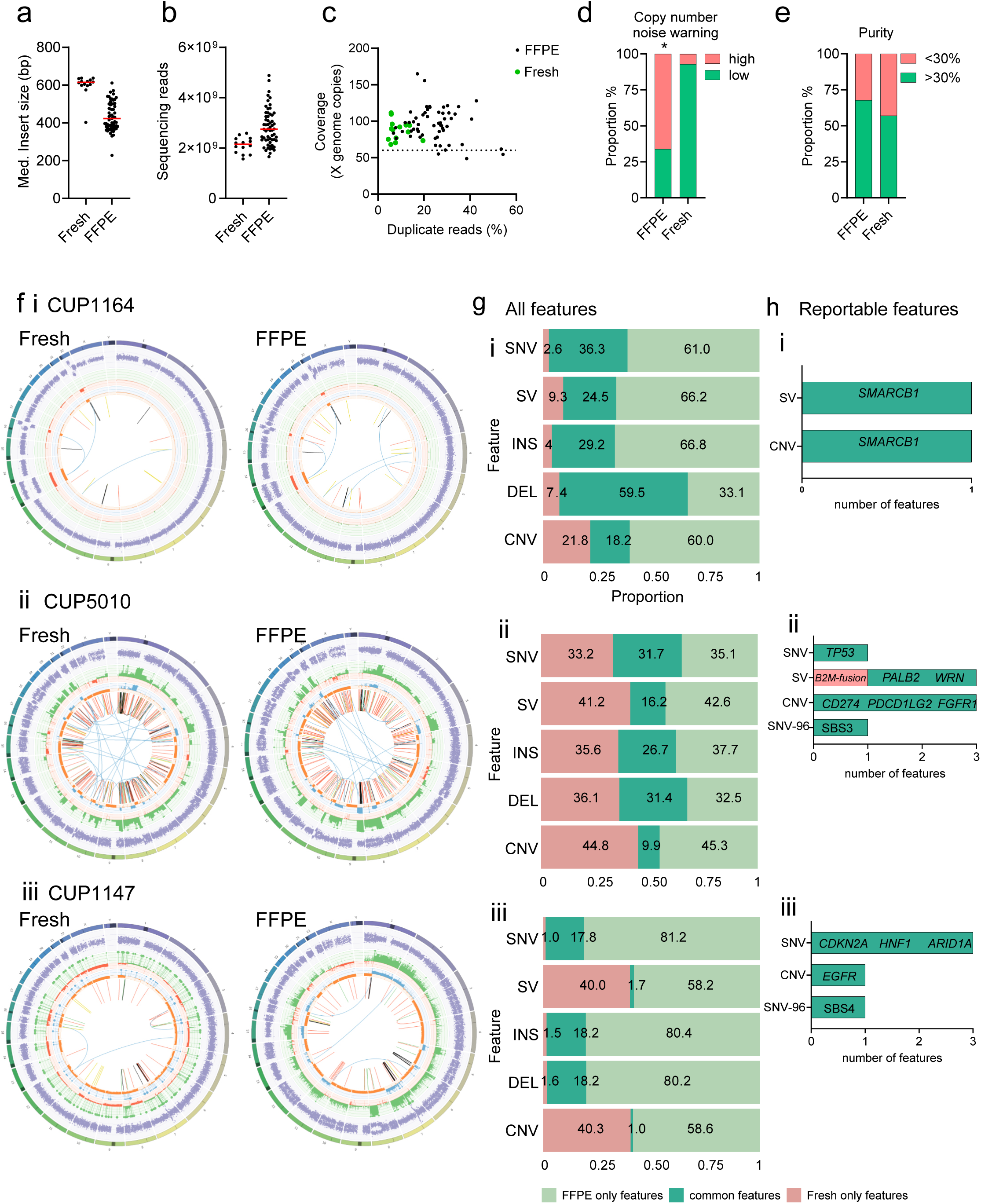
Feasibility of WGTS for CUP using FFPE specimens. **(a)** Median DNA fragment insert size by sample type and **(b)** number of raw sequence reads generated for 14 fresh and 59 FFPE samples. **(c)** Scatter plot of sequencing coverage against number of duplicate reads detected for 14 fresh and 59 FFPE samples. **(d)** Proportion of samples with a copy-number noise warning for fresh and FFPE samples in this study. *Chi-square χ^2^(1, N = 73) = 15.88, *P<0.*0001. **(e)** Proportion of samples with a tumour purity above and below 30% as estimated by PURPLE (see methods). **(f: i-iii)** Circos plots of matched WGS data from fresh and FFPE specimens for three CUP tumours (CUP1164, CUP5010 and CUP1147). From outside-in: Chromosome number, and B-allele frequency, outward facing peaks define regions of copy number gains/amplification: green (monoallelic) and blue (biallelic), inward facing peaks define regions of chromosome loss: red (monoallelic) and orange (biallelic) and the innermost circle shows gene structural rearrangements. **(g: i-iii)** Bar graphs showing distribution of overlapping features across each of the three paired fresh-FFPE samples. **(h: i-iii)** Number of ACMG/AMP/CAP consensus reported features captured and overlapping between fresh and FFPE samples.

Consistent with a prior study^17^, copy-number variation (CNV) noise was greater for FFPE samples in our series, reflected by the number of cases flagged with a copy-number noise warning (PURPLE tool, see methods) (Fig. 2d, *Chi-square χ^2^(1, N = 73) = 15.88, *P*< 0.0001, Supplementary data 2). A slightly higher tumour purity was observed for FFPE, most likely because these tissue samples underwent histological review and dissection of tumour regions prior to nucleic acid extraction, which was not done for fresh samples. By contrast, 6/14 (43%) of fresh tissue cases had low tumour purity (<30%), compared to 19/59 (32%) FFPE cases (Fig. 2e, Supplementary data 2).

Matched FFPE and fresh tissue biopsies were available for three patients enabling a direct comparison of WGTS data quality and reportable features. While the somatic profile was similar between the paired data (Fig. 2f), many discordant single-nucleotide variants (SNV) insertions (Ins), deletions (Del) and structural variants (SV) were detected, which was especially evident in two cases (CUP1164 and CUP1147, Fig. 2g). However, considering only reportable features (Fig. 2h) there was only one additional SV event found in one fresh tissue sample (CUP5010) compared to paired FFPE data, involving a loss of function *B2M*-*SATB2* fusion – a potential biomarker of resistance to immune checkpoint blockade therapy^19^. Tumour heterogeneity and independent sampling of fresh and FFPE samples may also explain discordant features between paired fresh and FFPE data.

In summary, despite FFPE tissue being inferior to fresh tissue and WGTS data quality being highly variable between cases, complete analytical failure using FFPE tissues was rare with 59/61 (97%) cases being suitable for clinical reporting.

### WGTS is superior to panel sequencing for the detection of reportable features

The availability of paired cancer panel and WGTS data enabled a direct comparison of the reportable features in 71/73 (97%) cases. Small somatic variants (SNVs & Insertions/deletions, Indels), CNVs and SVs were curated independently for WGTS and panel sequencing platforms. As curation protocols varied between platforms and over time, variant annotation for all data was retrospectively and centrally reviewed to enable a direct comparison of therapeutic and diagnostic biomarkers according to standardized AMP/ASCO/CAP consensus guidelines (see Methods) (Fig 3a).

**Figure 3.**
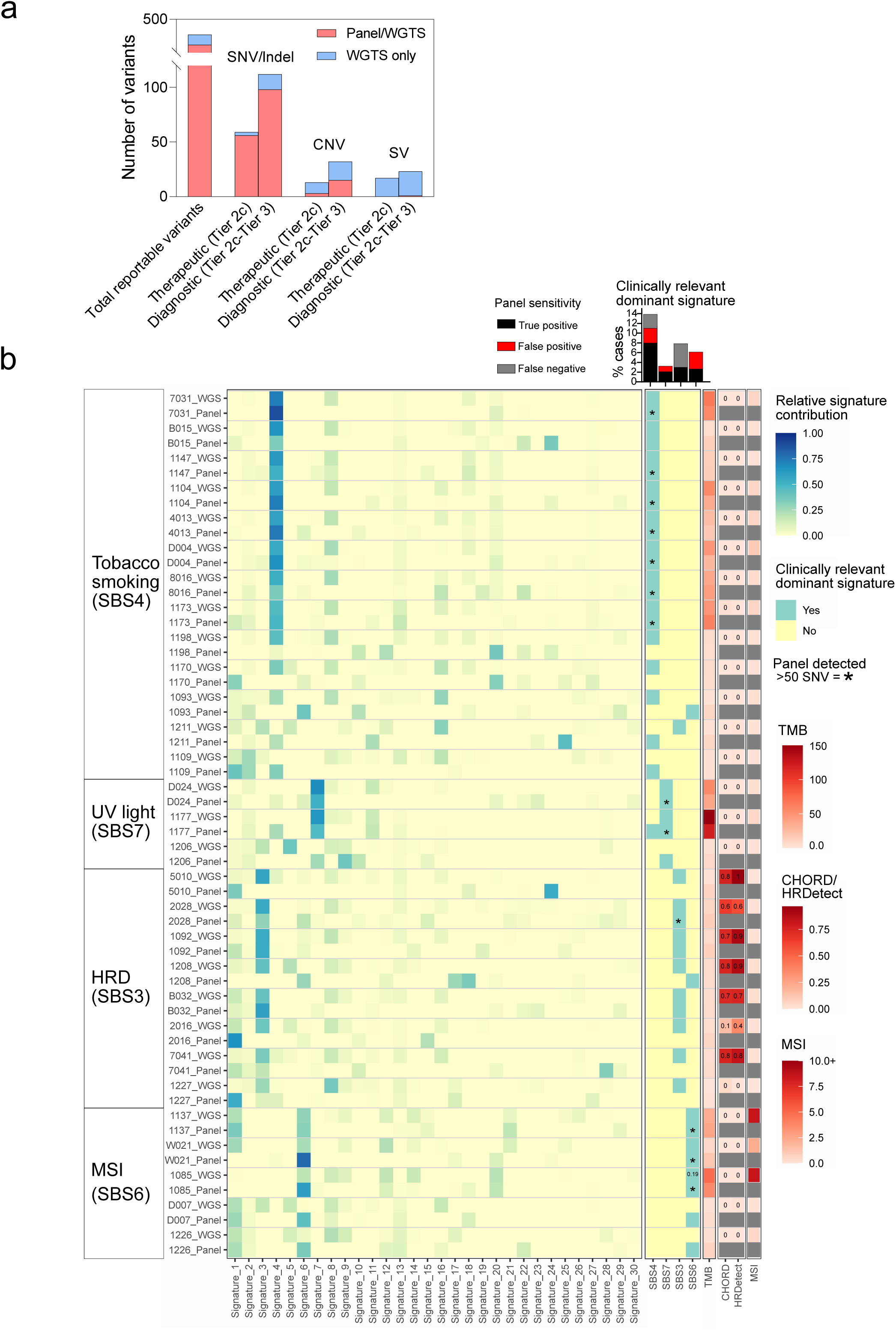
Detection of therapeutic and diagnostic mutational features in CUP using WGTS. **(a)** Number of therapeutic and diagnostic variants in CUP cases detected in both matched panel and WGTS or WGTS only assays. **(b)** Schematic showing comparison of relative SNV-96 mutational signatures detected in WGS with matched panel cases (COSMICv2). Percentage of SBS4, SBS7, SBS3 and SBS6 true positive, false positive and false negative cases scores are quantitated above. Asterisked (*) cases represent cases with greater than 50 SNV detected by panel. HRD (HRDetect and CHORD) and MSI (MSIseq) scores from WGS data are presented alongside for each case.

With respect to curated SNVs and small Indels, there was 88% (181/206) concordance between panel and WGTS data, but all variants detected by panel sequencing were found by WGS (Supplementary data 4). SNV/Indel somatic alterations missed by panel testing were likely explained by regions of interest not captured by the panel sequencing, such as the *TERT* promoter (Supplementary data 4). Detection of SVs and CNVs was a major advantage of WGTS over panel, with most CNVs (37/58, 63%) and nearly all SVs (54/55, 98%) detected only by WGTS.

In total, WGTS identified 89 therapeutic and 107 diagnostic variants in 65/73 (89%) tumours, whereas panel testing detected 58 therapeutic and 81 diagnostic variants in 41/71 (58%) of tumours (Fig. 3a).

### WGS improves detection of diagnostic and therapeutic mutation signatures

Somatic single-base substitution (SBS) signatures can be of high diagnostic and therapeutic value^20^. With respect to diagnostic signatures, WGS detected a dominant SBS4 (tobacco smoking) in 11/73 (15%) and SBS7 (ultra-violet light) in 2/73 (3%) cases, consistent with lung and skin cancer aetiology, respectively. (Fig. 3b). Comparatively, while panel sequencing could detect SBS7 in the two presumed skin-cancer CUPs, it could only detect SBS4 in 8/11 (72%) presumed lung-CUPs (Fig. 3b). Furthermore, as there was propensity for false-positive detection of dominant mutational signatures using panel in tumours with a low mutation burden, we set a minimum threshold of 50 mutations, which reduced the sensitivity for SBS4 detection by panel to only 7/11 (64%) cases.

Homologous recombination deficiency (HRD) can predict response to platinum chemotherapy or Poly-ADP-ribose polymerase (PARP) inhibition^13,21,22^. Two algorithmic HRD-prediction tools (CHORD^23^ and HRDetect^24^) were applied to WGS data. CHORD/HRDetect-positivity was detected in six cases. (Fig. 3b). Importantly, CHORD/HRDetect-positivity corresponded with somatic or germline mutations in HRD-related genes in four cases, and two cases (CUP1092 and CUP1208) were also deemed HR-deficient in the absence of HR-gene mutations, that would have made these patients eligible for a clinical trial (such as IMPARP-HRD: NCT04985721) (Fig. 3b). HR-Detect and CHORD have not been validated using cancer panel data therefore this was not attempted. Dominant SBS3 is also associated with HR-deficient cancers^13,25^ and was detected in 8/73 (11%) CUPs by WGS and only three cases by panel, although this was reduced to only one case by panel when applying a minimum threshold for mutation number (≥50 mutations). Furthermore, it is known that SBS3 can lack specificity when used in isolation^26^, and this was demonstrated for two CUP cases (CUP1227 and CUP2016) that had dominant SBS3 but were otherwise deemed HR-proficient by CHORD/HR-Detect.

Microsatellite instability (MSI) and high tumour mutational burden (TMB) can also predict clinical response to immune checkpoint blocking antibodies^27–29^. An MSI caller was applied to WGS and TSO500 panel data (see methods), but not CCP panel given it was not designed for MSI detection (Fig. 3b). Three of 73 CUP cases (4%) had MSI-High ( by WGS and harboured additional supporting MSI features: e.g. MLH1 protein loss of expression in tumour cells by immunohistochemistry and high TMB (>10 mut/Mb)^13^ (Fig 3b). Interestingly, in these three MSI-High CUPs, the TSO500 MSI score (see methods) was below the threshold of MSI-High. Dominant SBS6 signature, which is a known feature of MSI, was detected by both panel and WGS, but this is not typically used for MSI detection, and again a dominant SBS6 signature had a propensity to be falsely called by panel (e.g. D007 and 1226) when less than 50 mutations were detected (Fig. 3b).

Overall, TMB was highly correlated between WGS and with panel sequencing (CCP vs WGS, r= 0.96; TSO500 vs WGS, r=0.97) (Supplementary Fig. 1a, Supplementary data 3). Considering the TMB threshold for FDA-approved immune checkpoint treatments (≥10 muts/Mb), 17/71 (24%) of tumours had high TMB by panel but only 12 of these by WGS. This was generally consistent with panel sequencing generating a higher TMB estimate in 54/71 (75%) or cases (Fig.3b, supplementary Fig. 1b, supplementary data 3).

As expected, WGS was therefore more sensitive for the detection of genome-wide mutational signatures of diagnostic and therapeutic importance, which included tobacco smoking, HRD and MSI signatures, whereas a high TMB score was more often given by panel sequencing.

### Mutation features can inform a clinicopathological diagnosis

We previously showed that genomic features including somatic driver mutations, mutational signatures and oncoviral DNA can be useful to support a TOO diagnosis^14^. Diagnostically informative features were determined based on an enrichment of gene specific mutations in cancer types using AACR Project Genie data as a reference (Supplementary Fig. 2). Using WGTS and panel data for the current series we again considered diagnostic mutation features to inform histopathology review, which included use of the available immunohistochemistry and clinical data (Fig. 4a). Among the 58 WGTS cases initially considered clinicopathology-unresolved before genomic testing 31/58 (54%) could now be assigned a single likely TOO diagnosis based on the genomic evidence (Fig. 4b-c). Furthermore, a narrowed anatomical location was possible for an additional 7/58 (12%) cases. For example, detection of HPV16/18 (HPV+) indicated a primary site of either anogenital or head and neck regions, but there was insufficient evidence to resolve what region.

**Figure 4.**
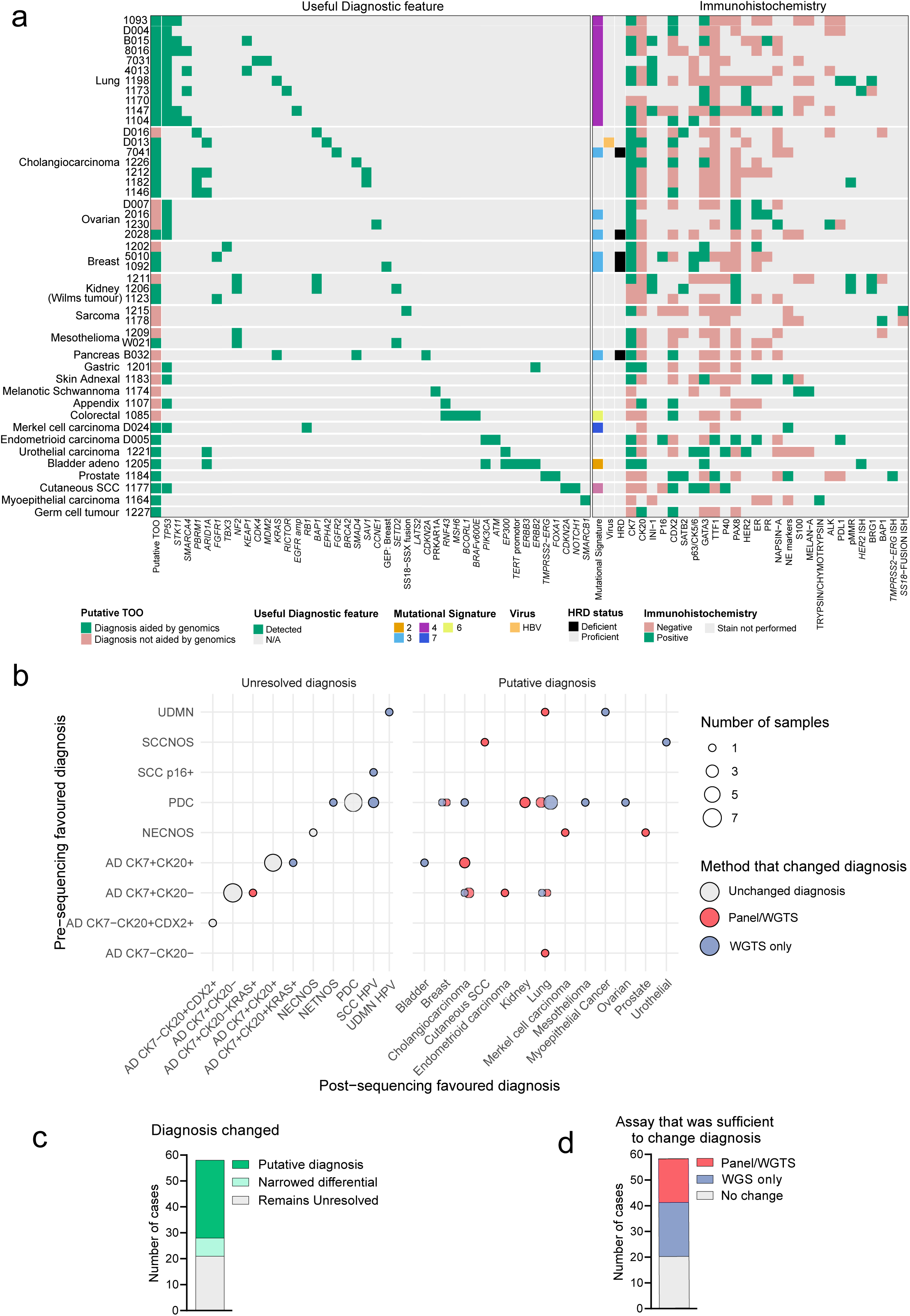
WGTS features aid clinicopathology work up of CUP. **(a)** Schematic of resolved CUPs grouped by favoured cancer type showing diagnostic molecular features in addition to immunohistochemical features. Diagnoses that were aided by genomics information are indicated in green (n=31) and those that were not are indicated in red (n=15). Molecular and IHC features that were detected or stained positive, respectively, are coloured green and those that were negative are coloured red. **(b)** Bubble chart showing MSKCC OncoTree cancer classification of clinicopathology-unresolved CUPs before and after cancer panel sequencing and WGTS (n=58/73). CUP tumours are grouped as clinicopathology unresolved or assigned a putative TOO following genomics profiling and are colour coded by the method that changed their diagnosis. **(c)** Number of clinicopathology unresolved cases that had a diagnosis changed because of additional genomics findings. **(d)** Number of resolved cases aided by panel only findings or that required additional WGTS features to resolve a TOO.

In summary, interpretation of panel findings alone was sufficient to aid primary TOO diagnosis in 19/58 (33%) cases that could not be assigned a TOO based on initial pathology review, but this increased to 31/58 (54%) cases when considering the WGTS findings (Fig. 4d).

### Algorithmic tissue of origin prediction complements data curation and clinicopathology

CUPPA uses orthogonal DNA features from WGS to classify unknown tumours into 36 defined cancer classes^11^. For the current study we extended CUPPA to include WTS data, enabling independent RNA classification as well as a combined “DNA+RNA” prediction (Fig. 5a and see methods). We tested the accuracy of the CUPPA DNA+RNA by leave-one-out cross-validation (LOOCV) using a training dataset of 6106 primary and metastatic tumours of known origin (PCAWG & HMF datasets) (Supplementary fig. 3).

**Figure 5.**
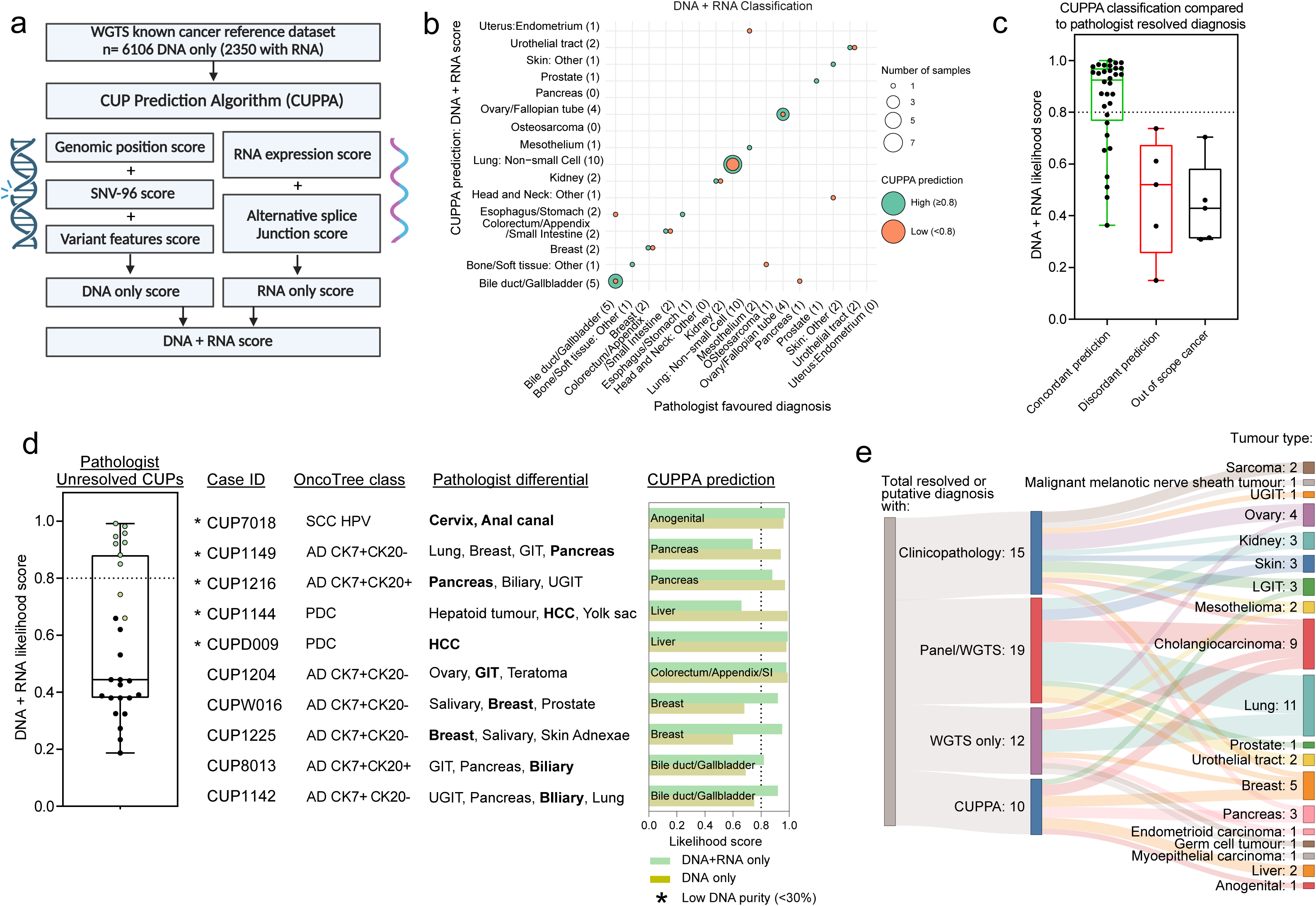
Application of a tissue of origin classifier to aid a CUP diagnosis. **(a)** Flow chart of the CUP prediction algorithm (CUPPA) that classifies CUP samples toward 1 of 36 defined cancer types by using a large reference dataset of known cancers. CUPPA (version 1.4) calculates 5 orthogonal variant feature scores for each class: 3 that combine into an overall DNA score and 2 that combine into an overall RNA score. Combined DNA + RNA score is the final reportable prediction for tissue with greater than 30% tumor purity. **(b)** Confusion matrix of DNA + RNA combined prediction scores (n=69 CUPs) against a pathologist’s favoured diagnosis, colour coded by high (≥0.8) or low (<0.8) likelihood. **(c)** Box plot showing combined DNA + RNA prediction scores for CUPPA using classification toward a single site tissue of origin prediction, categorized by concordance with a genomics informed, but CUPPA-blinded, pathology review. Cases with a favoured origin not represented in CUPPA training data were separated into a third group: out of scope cancer. **(d)** Box plot of all genomics-informed pathologist-unresolved CUP and schematic of ten of these CUP samples that were resolved with CUPPA and their high-likelihood (DNA+RNA or DNA only) predicted CUPPA class **(e)** Sankey plot of CUPs with a resolved TOO flowing toward the method by which they were resolved and cancer type.

The DNA+RNA classifier had a LOOCV-accuracy of 91.3%, which was superior to using DNA-only or RNA-only classifiers that had LOOCV-accuracies of 86.6% and 82.7%, respectively. The accuracy of the DNA+RNA classifier improved to 98.2% by setting a high-likelihood cut-point (0.8), with high-likelihood predictions representing 83.5% of all training set samples by LOOCV (Supplementary fig. 3b-c).

We next applied CUPPA to WGTS data from our CUP patient series. As WTS data was unavailable in four cases the CUPPA DNA-only classifier was used instead. We also applied the CUPPA DNA-only classifier to cases where tumour purity was <30% (inferred from WGS) reasoning that overrepresentation of RNA from adjacent normal tissue may confound gene-expression TOO classification. A high-likelihood CUPPA prediction was made for 41/73 (56%) CUP tumours. (Fig 5b-c, Supplementary Fig. 4a,c). The two most recurrent cancer type predictions were non-small cell lung cancer (NSCLC) (11/73 cases, 15%) and cholangiocarcinoma (7/73 cases, 9.5%). Importantly, among lung-CUP cases we observed that the DNA-only classifier was superior to RNA-only classification alone with respect to the proportion of high-confidence predictions made (Supplementary fig. 4e). Similar to our previous study using a Nanostring gene-expression classifier, this suggests many lung-CUPs have an atypical transcriptional profile^14^.

CUPPA predictions were next compared to the pathologist’s favoured primary TOO from the genomics-informed pathology review (Fig. 5b-c). Importantly, pathology review was done blinded to CUPPA results (Fig 5c, supplementary data 1). Notably, all cases with high-likelihood CUPPA predictions were concordant with genomics-informed pathology review (Fig. 5c, supplementary Fig. 4c-d). Conversely, among cases with low likelihood CUPPA predictions, nine were still concordant with pathology review, five were incorrectly classified, while five represented rare cancers and not represented in the training data (e.g. CUP1174, a malignant melanotic nerve sheath tumour, Fig 5b-c), (Fig 5c, Supplementary fig. 4f). Ten CUP tumours not resolved conclusively by genomics-informed pathology review had high-likelihood CUPPA predictions. These CUPPA predictions were still within the pathologist’s favoured diagnostic differential and therefore plausibly correct (Fig. 5d). When considering all cases that were either classified with high-likelihood CUPPA predictions and/or were resolved by pathology review, 56/73 (77%) tumours were assigned a single site primary TOO (Fig. 5e).

### Improvement in treatment options using WGTS

We next assessed the treatment options considering both therapeutically actionable mutations detected and the predicted TOO of a patient’s cancer, because TOO was often needed for drug access according to trial eligibility or standard-of-care treatment. Again, treatment options were assessed for the entire cohort retrospectively at single time point based on open phase I-III clinical trials in Australia and approved SOC treatments (Fig. 6a). As CUP1209 had 2 synchronous primary tumours (Mesothelioma and Ad CK7+ CK20-), this patient would not be considered for targeted therapy so was excluded from this analysis.

**Fig 6.**
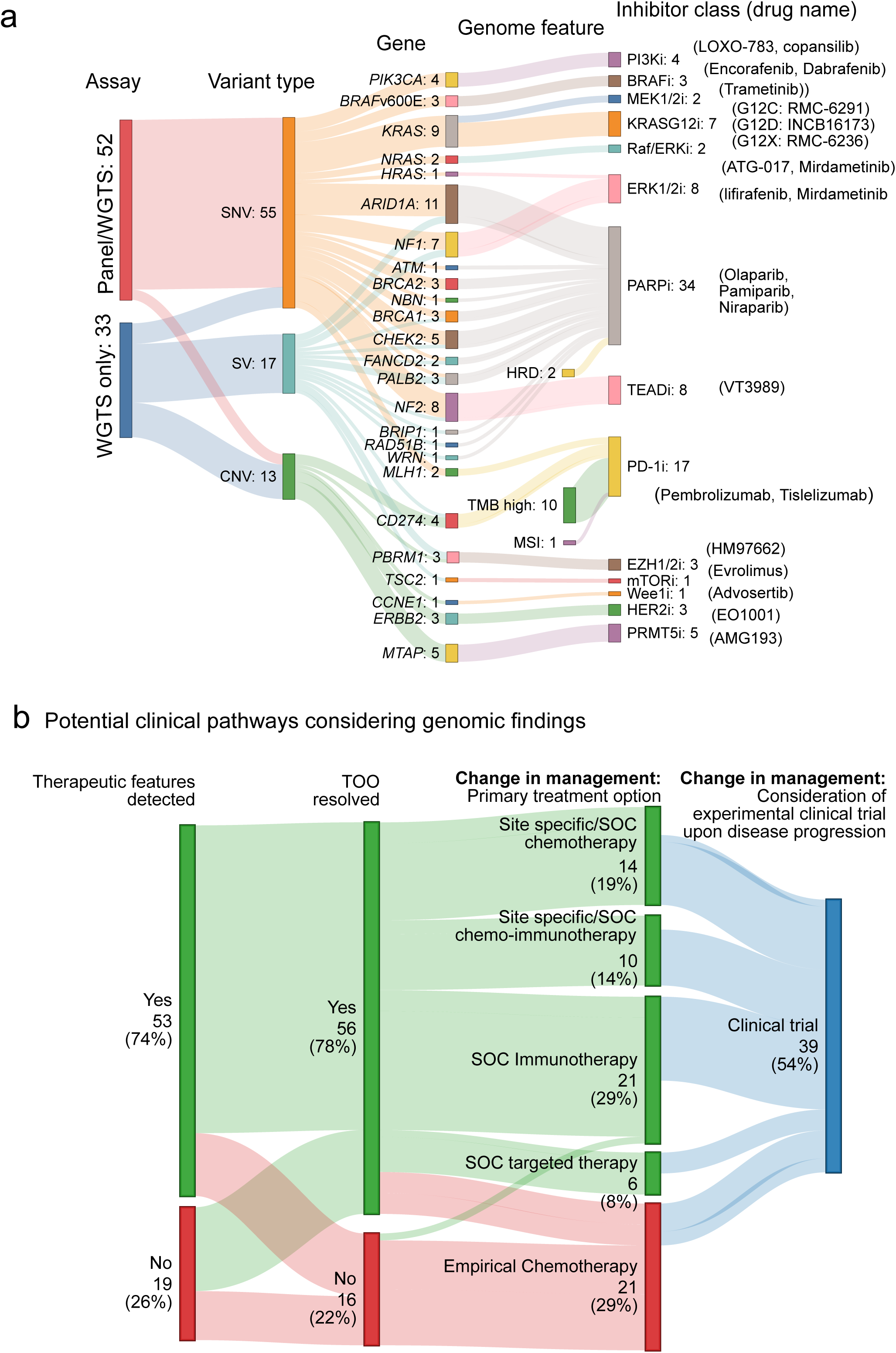
Using therapeutic biomarkers and TOO predictions to change clinical management. **(a)** Sankey plot showing all therapeutic features captured in matched panel and WGTS or only WGTS assays for 73 CUP samples, separated by variant type and gene or genome wide feature, flowing to eligible inhibitor (class and specific drug names). **(b)** Sankey plot of 72 CUP patients showing the number and proportion that had one or more therapeutic biomarkers detected, whether a putative TOO was determined and whether a SOC treatment and/or clinical trial could be considered to gain access to appropriate treatment.

WGTS identified one or more therapeutic biomarkers in 53/72 (74%) patients, while a putative diagnosis was found in a partially overlapping group. The combined diagnostic and treatment-related information would have directed SOC treatment for 51/72 (71%) patients and/or consideration for clinical trial in 39/72 (54%). (Fig. 6b, supplementary data 5). In contrast, panel sequencing detected therapeutic features in 49/71 (69%) but fewer patients would have had access to SOC therapy (35/71, 49%) or clinical trials (29/71, 41%) as a TOO diagnosis was not made. (Supplementary fig. 5, supplementary data 5).

Importantly, resolving TOO diagnosis alone would have provided access to SOC immunotherapy in 27/72 (37.5%) patients, according to Australian treatment guidelines for metastatic NSCLC (10), cholangiocarcinoma (8), cutaneous SCC (1), Merkel cell carcinoma (1), bladder cancer (1), clear-cell renal cell carcinoma (2), unresectable mesothelioma (1), hepatocellular carcinoma (2) and cervical cancer (1) (Fig. 6b). These results highlight the growing use of immunotherapy across several cancer types where a TOO diagnosis in CUP is necessary for SOC drug access.

In summary, WGTS indicated a potential treatment option for 79% of patients considering either SOC or clinical trial, compared to 62% of patients considering only the panel data.

### Assessing the utility of ctDNA-WGS for TOO prediction

To explore the application of cell-free DNA (cfDNA) for WGS as a substitute to tissue testing, we collected blood-plasma from 144 CUP patients (Fig. 7A). The median yield of cfDNA from CUP cases was 18.34 ng/mL (range 2.8-834 ng/mL) (Fig. 7B). Of these cases, 76/144 (52.7%) yielded a minimum amount of cfDNA (20 ng) to generate libraries suitable for WGS. (Fig 7A).

**Fig 7.**
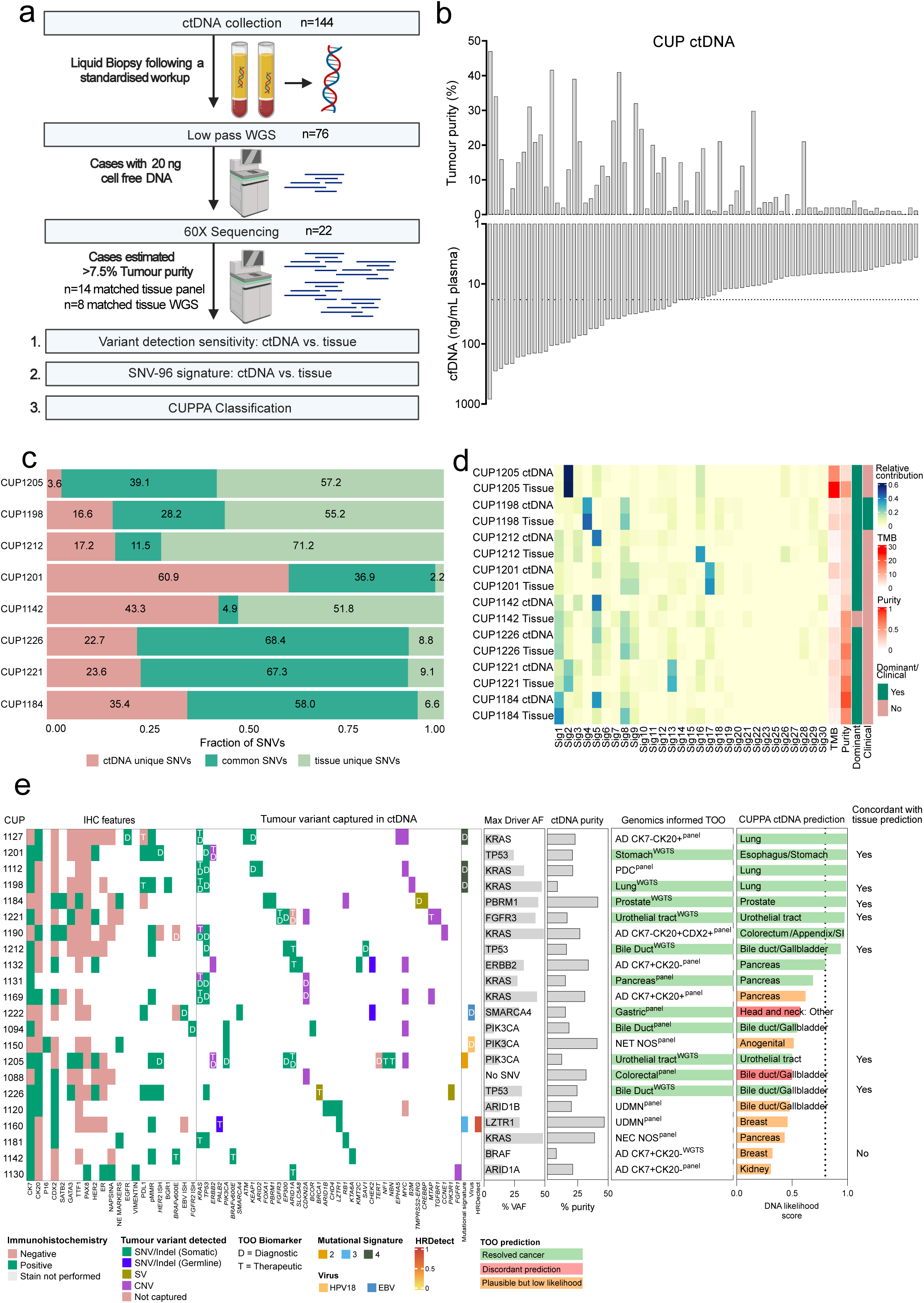
Utility of liquid biopsy derived ctDNA for WGS in CUP. **(a)** Flow chart diagram showing workflow for WGS of liquid biopsy samples from CUP patients. **(b)** Cell free DNA yield per mL of plasma and tumour purity estimates of cfDNA samples as calculated by the ichorCNA method across 76 patients. **(c)** Bar graph showing distribution of unique or common SNVs in 8 cases with matched ctDNA and tissue WGS. **(d)** SNV-96 mutational signatures (COSMICv2) across 8 cases with matched ctDNA and tissue WGS. Tumour purity (ichorCNA estimated), TMB, and the presence or absence of a dominant signature (greater than 20% abundance) or clinically relevant signature is shown alongside for each sample. **(e)** Schematic of the 22 CUP-ctDNA cases showing immunohistochemistry profiles of biopsied tissue alongside known genomic features from the tumour that were detected in ctDNA-WGS data. Percentage known driver variant allele frequency (%VAF) and ichorCNA estimated purity (%purity) for each sample is shown alongside. CUPPA predictions from ctDNA-WGS data are shown alongside for each case, as well as whether the tissue and ctDNA predictions are concordant with each other.

We first applied low-coverage whole genome sequencing (lcWGS) to determine the relative fraction of circulating tumour DNA (ctDNA) in blood plasma (0.6x coverage, ichorCNA method^30^). The estimated tumour fraction across the cohort ranged from 1-47%, and a median of 7% across 76 samples (Fig. 7B). Twenty-two CUP cases with ≥7.5% ctDNA fraction were subjected to deeper sequencing (median 118x coverage). (Fig. 7A). Of these cases, 8/22 (36.3%) had matched tissue-based WGS data available, while 14/22 (63.6%) only had cancer panel data available.

We next processed the cfDNA-WGS data through our standard clinical reporting pipeline to compare SNV detection and the similarity of SBS mutational patterns (Fig 7C-D). The average percentage of common SNVs detected in cases with paired WGS data (8/22) for tissue and ctDNA was 39.3% (range 4.9-68.4%). For the cfDNA-WGS samples with only matched CCP or TSO500 panel data (14/22), we identified the same tissue-reported mutations, in all cases, confirming the ctDNA had originated from same tumour sequenced. (Supplementary data 6).

The same dominant SBS signatures were found in matched ctDNA and tissue WGS data in 6/8 (75%) cases, which included signatures of diagnostic utility. (Fig. 7D). For example, a dominant SBS4 (tobacco smoking) was identified in a suspected lung-CUP case (CUP1198). We also detected SBS17 in CUP1201, a resolved oesophageal cancer, and this is a signature with unknown aetiology that has been reported at high frequency in oesophageal tumours^25,31^. (Fig. 7D).

Finally, we applied CUPPA DNA-only classification to the 22 cfDNA-WGS cases to compare CUPPA predictions using tissue-based WGS as well as cancer type based on clinicopathology review (Fig. 7E). CUPPA classified 9/22 (41%) cases with high-likelihood prediction, and these were all concordant with the favoured TOO or within a likely diagnostic differential. Of the remaining 13 cases with low-likelihood prediction only two cases were discordant with the pathologist favoured diagnosis. Importantly, CUPPA using cfDNA-WGS made a high-likelihood prediction for four cases that were previously unresolved as only with panel data was available (CUP1127 & CUP1112: NSCLC, CUP1190: Colorectum/Appendix/SI and CUP1132: Pancreas). Altogether, this demonstrates the potential to apply WGS to cell-free DNA and to make confident predictions of cancer type where there is insufficient tissue for WGS analysis.

## Discussion

Genomic profiling is increasingly being used as part of clinical testing in CUP patients, supported by recent recommendation of tumour mutation profiling in international CUP guidelines^1,32^. Although gene-panel sequencing can detect many approved therapeutic targets, WGS increases the diagnostic yield for treatment-decision making^33^. In the current study, we have also shown that WGTS is feasible for real-world clinical samples and can be as sensitive as panel testing for mutation detecting, even when using FFPE samples. Our systematic comparison of targeted gene-panel and WGTS platforms confirmed that WGTS significantly increased the number of therapeutic and diagnostic features identified. Furthermore, algorithmic TOO prediction using WGTS data could assist in resolving the TOO of many CUP tumours. Importantly, we show this method may also be extended to use on high tumour-fraction cfDNA samples, thereby increasing accessibility of testing for CUP patients.

Several studies have described the feasibility of WGTS for paediatric and adult solid cancers, but few have supported the use of FFPE tissue specimens^34–40^. A pilot study of the 100,000 Genomes Project found WGS of FFPE nucleic acids was technically feasible in 52 cases. However, the data quality was adversely impacted by formalin fixation leading to increased copy-number noise, that could be improved by modifying the DNA reverse cross-linking protocol required for DNA extraction^17^. Nevertheless, in the current study we achieved a high WGTS success rate (97%) using FFPE samples when we applied DNA QC methods prior to library preparation. Good concordance of reportable features was observed between three paired fresh-FFPE cases and comparable results to cancer panel sequencing in 71 cases, where no loss of sensitivity was observed in panel captured regions. Although sequencing costs are higher for WGTS of FFPE compared to fresh samples, this may be partially offset by the additional cost of re-biopsy, while also reducing the patient morbidity associated with invasive biopsy procedure. Moreover, the collection and processing of fresh tissue samples remains logistically challenging for many centres, especially in regional parts of Australia, creating a potential disparity in test access between treatment centres.

We have demonstrated that the yield of reportable treatment-related and diagnostic findings was higher for WGTS compared to panel testing. A previous study used *in-silico* comparison of WGS, whole exome and multi-gene testing showing panel sequencing was sufficient for detecting most treatment biomarkers, but WGS could detect more treatment biomarkers required for clinical trials^41^. Similarly, we found an increase in potential clinical trial eligibility using WGS data, with 24% more patients eligible for access to SOC therapy and/or phase I-II clinical trials. Importantly, we found that large structural events (CNV, SV) constituted most of the additional diagnostic information found by WGTS. SVs can occasionally involve pathognomonic drivers, such as *TMPRSS2-ERG* in prostate cancer and *SS18-SSX* in synovial sarcoma, as highlighted in recent ESMO guidelines^1^. These events can also be detected by some commercial panels or histological methods like fluorescence in-situ hybridisation; however, non-recurrent SV events involving tumour suppressor genes are often missed by panel testing. For example, among CUP tumours profiled, *PBRM1* mutations and SVs were identified in four of seven CUPs thought to be intrahepatic cholangiocarcinomas by pathology review and confirmed by CUPPA prediction. *PBRM1* events can be found in ∼8%^42,43^ of cholangiocarcinomas and ∼39% renal cell carcinomas^44^, therefore both cancers may be considered; however, these tumour types have distinct histological appearances therefore highlighting the need to always interpret mutation data with available clinical and pathological information.

Mutational signatures are another DNA feature that can contribute to diagnosis where WGS was also superior. While SBS4 (tobacco smoking) and SBS7 (UV light damage) can both be detected by panel, these signatures can be missed due to a lower number of variants detected by panel sequencing. Aside from being of diagnostic value, mutational signatures are of high therapeutic importance, and we found WGS was superior not only for detecting HRD but also surprisingly MSI, although the patient numbers were small. Although targeted capture sequencing methods have been recently developed for detection of HRD and are used for cancer types such as ovarian (HRDscar^45^, Myriad Genetics, USA), breast and pancreatic cancer (HRDsig^46^, Foundation Medicine, USA) these tests would not typically be applied for CUP patients. For instance, we found HRD and a *BRCA2* mutation in a CUP tumour ultimately resolved as a cholangiocarcinoma. Although it may be plausible to detect HRD in a sequential manner to DNA panel testing, the competitive affordability of using targeted sequencing is eroded when there is a need to apply multiple panel tests.

WGTS was able to resolve a likely TOO in most CUP patients using a combination of curated genomic features to inform clinicopathology review and CUPPA. We previously reported that panel testing can assist TOO diagnosis in 31% of cases^14^. In an independent cohort of 73 CUPs, we now show that WGTS can aid in resolving up to 77% of cases. CUPPA alone made high-confidence predictions in 56% of cases, and all predictions were either concordant with the pathologist’s opinion or within their diagnostic differential. Similarly, Schipper *et al.*^11^ used CUPPA (DNA only) for 72 CUP tumours to assign a putative diagnosis in 64% of cases. While deep-learning classification has also been applied to panel data, such as the recently described OncoNPC^12^ method, which made high-confidence classifications in only 41% of CUP cases. Importantly, CUPPA accuracy was improved by combining DNA and RNA evidence, which was especially important for cancer types such as NSCLC. Similar to our previous study comparing DNA and RNA panel tests, gene-expression similarity alone is unreliable for classification of lung-CUP tumours that have atypical expression patterns, including absence of IHC staining for the classic lung marker TTF1^14^. These tumours can frequently harbour disruptive mutations in the SWI-SNF chromatin modifier *SMARCA4* (BRG1), and we found *SMARCA4* mutations in four of 11 lung-CUPs in the current series. Importantly, there was good concordance between CUPPA prediction and genomics-informed pathology review of our CUP series. While this validates CUPPA, it also validates the approach of using curated genomic features to assist in a pathology assessment. Importantly, using genomic features to inform pathology review was especially important for rare cancer types that were out of scope of the current CUPPA training set.

Despite the potential to use FFPE samples for WGTS, access from pathology archives can still be slow and lead to extensive delays in testing^47^, while in some cases tissue samples may be completely exhausted from immunohistochemistry testing. CUP patients are known to have high amounts of cfDNA in their blood and panel testing has been shown to detect reportable findings in up to 80% of patients^48^. We found just over half of our CUP patient series had sufficient cfDNA from a 20 mL blood draw for WGS and that the tumour fraction in cfDNA was high enough to apply standard WGS methods in a third of patients, including application of the CUPPA method. Despite our data showing concordance between CUPPA predictions using matched cfDNA and tissues, further improvements may be likely to increase the sensitivity of the approach, perhaps through increasing WGS sequence coverage or modifications to analytical pipelines adapting to low tumour fraction cfDNA samples. We note that TOO classification is also possible using DNA methylation analysis^9,49–51^ therefore combining DNA methylation with mutation detection for cfDNA analysis is likely to improve classification.

The adoption of WGTS into routine diagnostic workup is likely to increase with reduced cost of sequencing. We have shown the clinical validity of this technology in the work-up of CUP patients using real-world clinical samples, increasing the sensitivity for detecting therapeutic targets and identifying diagnostic features that can aid a TOO diagnosis.

Further improvements will increase the sensitivity of the CUPPA method to improve TOO classification, especially among rare cancers as well as when using cfDNA samples. Although the diagnostic and therapeutic applications of WGTS for CUP is apparent, wider acceptance of such genomic tests for cancer diagnosis and treatment decision-making requires independent prospective validation to determine the clinical impact.

## Acknowledgements

We wish to thank SUPER study coordinators at participating hospitals for facilitating collection of biospecimen and data for the study. We wish to acknowledge the patients who have contributed to this study and the CUP consumer steering committee: Cindy Bryant (chair), Kym Sheehan, Christine Bradfield, Clare Brophy, Dale Witton, and Frank Stoss. The study was supported by funding from Australian Health Genome Alliance (AGHA) and National Health and Medical Research council (NHMRC) (1113531) and the Medical Research Future Fund (MRFF, GHFMCDI000003). RWT was supported by funding from the Victorian Cancer Agency (TP828750). The Westmead, Blacktown, and Nepean study sites were supported by the Cancer Institute NSW 11/TRC/1-06, 15/TRC/1-01, and 15/RIG/1-16. We acknowledge the TAGC clinical genomics platform group for their valuable contributions.

## Author contributions

RWT and LM conceived the study. RJR and AP performed the analysis. RJR and RD drafted the figures and tables. OWJP undertook histopathology review of all CUP cases. TS and LM reviewed the clinical data and RJR, CamM, WV and TS retrospectively reviewed clinical trials eligibility based on biomarker data. CF, CW, KF and SW collected clinical data for the study. HW, ADF, NW, CSK, BG, CSt, MS, IMC, GR, MW and NK screened patients for eligibility. CSh and PP developed the CUPPA classifier. CBM, JV, WZ, SN, AFe, HX and SF curated mutation profiling data. RD, SK and AFl performed the bioinformatic analysis. RJR and RT co-wrote the manuscript and all authors critically reviewed the manuscript. SG and OH oversaw the clinical genomic profiling and bioinformatic cohort profiling for this study. PS, DB, LM and RT were the principal investigators and obtained research funding to support the study.

## Methods

### CUP Clinicopathological review

All CUP patients were recruited to the SUPER study from 11 Australian sites between 02/10/2017 and 20/10/2021 with informed consent under an approved protocol at Peter MacCallum Cancer Centre (PMCC) human research ethics committee (HREC protocol: 13/62). No patient identifying information has been included in this study and all patients are referenced with anonymous identifiers, where identities are only known to the study team. Blinded histopathology review for the tissue WGS set (n=72 cases and n=73 distinct tumours) was performed by a single pathologist (OWJP) (supplementary data 1). Cases were assigned a favoured TOO upon registration to the study and then assigned a revised favoured TOO based on a curated panel and WGTS report (supplementary data 1). The diagnosis was reassessed retrospectively as clinicopathology-unresolved or putative diagnosis for all cases. This was done unblinded to genomics curated reports but blinded to the CUPPA result in all cases (n=87, tissue and ctDNA). When classification could not be reached, a modified version of the Memorial Sloan Kettering Cancer Centre (MSKCC) Oncotree classification criteria^52^ for CUPs was used to subclassify malignancies, and these were: undifferentiated malignant neoplasms (UDMN); poorly differentiated carcinoma (PDC); adenocarcinoma, not otherwise specified (ADNOS); neuroendocrine tumours, not otherwise specified (NETNOS); neuroendocrine carcinomas, not otherwise specified (NECNOS); and squamous cell carcinomas, not otherwise specified (SCCNOS). ADNOS were further subdivided based on cytokeratin 7 (CK7) and cytokeratin 20 (CK20) IHC staining; where CK7 was negative and CK20 had positive staining, caudal type homeobox 2 (CDX2) positivity was annotated. SCCNOS were subclassified based on p16INK4A (p16) IHC staining positivity.

### Tissue and blood collection and nucleic acid extraction

Where possible fresh tissue biopsy specimens were collected and stored in “RNAlater” (Thermo Fisher, USA, cat. #AM7020) for 24 hours before DNA and RNA extraction. Otherwise, an archived (FFPE) tissue specimen was used. Representative sections were reviewed by a pathologist and tumour regions macro-dissected before nucleic acid extraction. Only regions > 30% tumour cellularity were selected for WGTS. DNA and RNA extraction was done using the AllPrep DNA/RNA FFPE kit (QIAgen, USA, #80234). Alongside, for each recruited patient, a 25 mL whole blood sample was collected in EDTA tubes (5 mL) or Streck DNA Blood Collection Tubes (Streck, USA). Germline DNA was exclusive derived from whole blood in EDTA tubes. Blood plasma was derived only from streck tubes and stored at -80°C as 1 mL aliquots before cell-free DNA extraction with QIAamp circulating nucleic acid kit (QIAgen, USA, cat. #55114).

### Cancer gene panel DNA sequencing

All cancer panel sequencing was performed by the medical laboratory Nexomics, at the Peter MacCallum Cancer Centre. Twenty-seven samples were sequenced with a custom cancer panel (CCP) targeting 386 genes (Agilent SureSelect), involving sequencing of both tumour and matched germline DNA, as previously described^53^. Briefly, libraries were prepared and enriched using SureSelect XT enrichment (Agilent) and indexed libraries were pooled and sequenced to a targeted exon coverage of 150X using 2×75 bp reads on an Illumina NextSeq500 instrument. Seqliner v0.7 (http://bioinformatics.petermac.org/seqliner/) was used to generate aligned reads against the hg38 human reference genome. Somatic variants were detected with Mutect v2.2 (https://github.com/broadinstitute/mutect) and filtered with GATK FilterMutectCalls v4.1.8.1 (https://github.com/broadinstitute/gatk). TMB estimation was described previously^14^ using the ensemble variant caller bcbio-nextgen (BCBio) cancer somatic variant calling pipeline (version 1.1.3a) (https://github.com/bcbio/bcbio-nextgen). Forty-six tumour samples were sequenced with the TruSight Oncology 500 panel (TSO500, Illumina) targeting 523 genes for DNA mutation detection and 55 cancer genes for RNA fusion and splice variant detection. TSO500 Libraries were prepared as per the manufacturer’s instructions and sequenced to a targeted mean exon coverage of 150x on an Illumina NextSeq500 instrument. Illumina Software TSO500 v2.0 Local App performed read alignment and variant calling against the hg37 human reference genome.

### Whole genome and transcriptome Sequencing

FFPE-derived DNA were assessed for WGTS suitability based on a modified *GAPDH* multiplex PCR assay that qualitatively estimates proportion of non-overlapping DNA fragments of between 100 bp to 400 bp in a given sample^18^. We modified this to include additional primer pairs to amplify 500, 600 and 800 bp fragments (Supplementary table 1), where scores of 1-8 were assigned to a sample depending on the largest amplifiable fragment (minimum “% Integrated Area” of 10% as visualised by TapeStation 4200 D1000 electropherogram). Samples were deemed suitable for library preparation when they scored at least 4/8, indicating minimum “% Integrated Area” of 10% was achieved for 400 bp length fragments as the largest fragment size. DNA Libraries were prepared using the Illumina TruSeq Nano library (Illumina, USA) method using 200 ng of DNA. All libraries were quality controlled using the TapeStation high sensitivity D5000 or D1000 ScreenTape (Agilent). Indexed libraries were pooled and sequenced aiming for a depth of 50x for normal and 100x for tumour using 150 bp paired reads on an Illumina Novaseq 6000 platform (Illumina, USA). Sequence reads were aligned to hg37 and processed using the Hartwig Medical Foundation (HMF) pipeline v5 (https://github.com/hartwigmedical/pipeline5). WGS data was re-processed for all cases using hg38 retrospectively by the Illumina DRAGEN pipeline (https://doi.org/10.1101/2023.03.23.534011) and umccrise workflow (https://github.com/umccr/umccrise). For WTS, RNA samples were subjected to ribosomal RNA depletion using the NEBNext rRNA Depletion kit (New England Biolabs) according to the manufacturer’s instructions. Approximately 100 million reads were generated per RNA sequencing library. WTS data was aligned using the STAR aligner^54^ (https://github.com/alexdobin/STAR) and feature counts were obtained as library-composition adjusted Transcripts Per Kilobase Million (adjTPM) using the Isofox algorithm (https://github.com/hartwigmedical/hmftools/tree/master/isofox). Fusions were confirmed in RNA-seq data using arriba (https://github.com/oicr-gsi/arriba).

TMB was calculated as outlined previously^55^. A ’high’ TMB was called if the TMB was over ≥10 mutations per Mb, and ’low’ if <10 mutations per Mb. MSI was estimated using the Personal Cancer Genome Report tool^56^ which utilizes a statistical MSI classifier from somatic mutation profiles that separate MSI-high from microsatellite stable tumours (https://rpubs.com/sigven/msi_classification_v3), a model based on MSIseq^57^. COSMIC V2 mutational signatures were assigned using ‘MutationalPatterns’ (v3.12.0)^58^. Dominant signatures were defined as those with greater than 20% of total somatic mutations within a sample.

Homologous recombination deficiency (HRD) was independently confirmed by HRDetect^24^ and CHORD^23^. HRDetect and CHORD consider mutational patterns (SNVs, InDels and SVs) characteristic of HR-deficient tumours. HRDetect grants a score from 0 to 1; tumour samples with a score > 0.7 are categorized as HR-deficient. CHORD classifies tumours into BRCA1-deficient and BRCA2-deficient categories; tumours with a combined probability of these HRD categories >0.5 are categorized as homologous recombination-deficient.

### Clinical and diagnostic features curation

A curation team at the Peter MacCallum Cancer Centre reviewed the findings from CCP and TSO500. For CCP, PathOS v.13^59^ (https://github.com/PapenfussLab/PathOS) was used to annotate variants, and filter for non-synonymous variants. Curation of germline variants was limited to 76 genes with evidence for cancer predisposition^60^. For TSO500, Clinical Genomics Workspace (CGW) from PierianDx was used to annotate, filter and report clinically relevant findings. The genomic events reported include single-nucleotide and multi-nucleotide variants, small Indels, CNVs, MSI and TMB. Dominant mutational signatures were calculated using MutationalPatterns v3.12.0 (https://github.com/AlexandrovLab/SigProfilerExtractor).

A curation team at the University of Melbourne Centre for Cancer Research reviewed all the findings for each case that were clinically reported. Curation of SV and CNV in WGS was limited to a custom list of 1246 cancer related genes, (https://github.com/umccr/umccrise/blob/master/workflow.md#key-cancer-genes) that were assembled using various sources. Manual inspection of DNA and RNA sequence reads was done to validate somatic variant calls using Integrative Genomics Viewer^61^.

Gene-fusions were supported by both WGS and WTS data. RNA-seq data also supported genome amplification or deletion of genes by comparing the normalized expression (z-score) of an altered gene in the CUP sample to tumours in a TCGA pan-cancer data set (https://github.com/umccr/RNAsum)

Regardless of sequencing approach, SNVs and Indels found in the tumour sample were prioritised into a four-tiered structure for clinical reporting based on actionable finding. This tiered structure adopted the joint consensus by AMP/ASCO/CAP on evidence based variant categorization^62^. We retrospectively reviewed and harmonized tumour variants based on therapeutic potential according to clinical trials available in January of 2024 and diagnostic potential according to significantly enriched variants within AACR Genie cohorts and/or WGS landscape papers as described previously^14^. Briefly, therapeutic variants belonged to AMP/ASCO/CAP consensus tier 2c, while diagnostic variants were AMP/ASCO/CAP consensus tier 2d if function was known or tier 3 if function was unknown but not/unlikely benign, therefore a variant could have been both therapeutic and diagnostic for a cancer type.

### Circulating tumour DNA extraction and sequencing

Cell-free DNA was extracted from plasma using the QIAamp circulating nucleic acid kit (QIAgen, cat. #55114), following the standard extraction protocol using a QIAvac vacuum manifold (QIAgen, cats. #19413, #19419 and #84020) with one modification. Our laboratory prepares a stock concentration of 1 µg/µL of carrier RNA for use in the plasma sample lysis step instead of 0.2 ng/µL. Up to 5 mL of plasma were used as input into a single extraction for a sample. Cell-free DNA was quantitated on an Agilent tapestation 4200, using cell-free DNA tapes (cat# 5067-5630). We determined sample concentration from a size limited range of 50-700 bp length DNA and cases were selected for library preparation if they had a minimum of 20 ng DNA. Whole genome libraries were prepared using the NEB NEXT Ultra II kit (New England Biolabs, #E7645). Indexed libraries were pooled and sequenced aiming for 1-2X coverage using paired 150 bp reads on the Illumina Novaseq 6000 platform (Illumina, USA). Tumour fraction was estimated using ichorCNA^30^ on each sample to determine suitability of libraries for deeper sequencing.

### CUP Prediction Algorithm (CUPPA) on tissue and cfDNA

The CUPPA tool, which is a Tissue of origin classifier that uses WGS and/or WTS data, was developed by Hartwig Medical foundation and described previously^11^. We used version 1.4 of CUPPA on eligible WGS and WTS data (https://github.com/hartwigmedical/hmftools/tree/master/cuppa). We utilized CUPPA DNA and RNA combined classifier outputs for final classification in tissue when RNA was available (69/73 cases), or only considered the DNA combined classification when RNA was unavailable (Supplementary data 1).

We considered “DNA only” classification for cfDNA samples where we had deeper coverage sequencing data (minimum 60x). We observed a pattern of false positive SNV calls in the 8 tissue and cfDNA matched WGS cases. To reduce the false positives in cfDNA samples, we ran Strelka2^63^ v2.9.2 (https://github.com/Illumina/strelka) and generated an ensemble of shared Strelka2 pass-filter calls and somatic SNVs from the HMF pipeline. In addition, we removed a customised panel of SNVs, which was constructed using somatic SNVs detected in two or more independent cfDNA samples but were absent in their matching tissues. HOTSPOT SNVs annotated by the HMF pipeline were not filtered.

### Figure generation

Figure generation was performed using R (v4.2.0), using tidyverse (v2.0.0), ComplexHeatmap (v2.18.0), patchwork (v.1.2.0). Flow charts were created with BioRender (https://biorender.com). Bar charts, scatter plots and statistical analyses were performed using GraphPad Prism version 10.0.0 for Windows, GraphPad Software, Boston, Massachusetts USA, (www.graphpad.com). Sankey plots were made using SankeyMATIC (https://sankeymatic.com/). All other graphics were created with Affinity Designer 2 (https://affinity.serif.com/en-us/designer/).

## Data availability

The datasets generated during and/or analysed during the current study are available in the European genome archive (EGA) repository, <u>[EGAS50000000452]</u>. Data is available from the EGA upon reasonable request. All analysed and curated data during this study are included in this published article (and its supplementary information files).

## Figure Legends

**Supplementary Figure 1.**
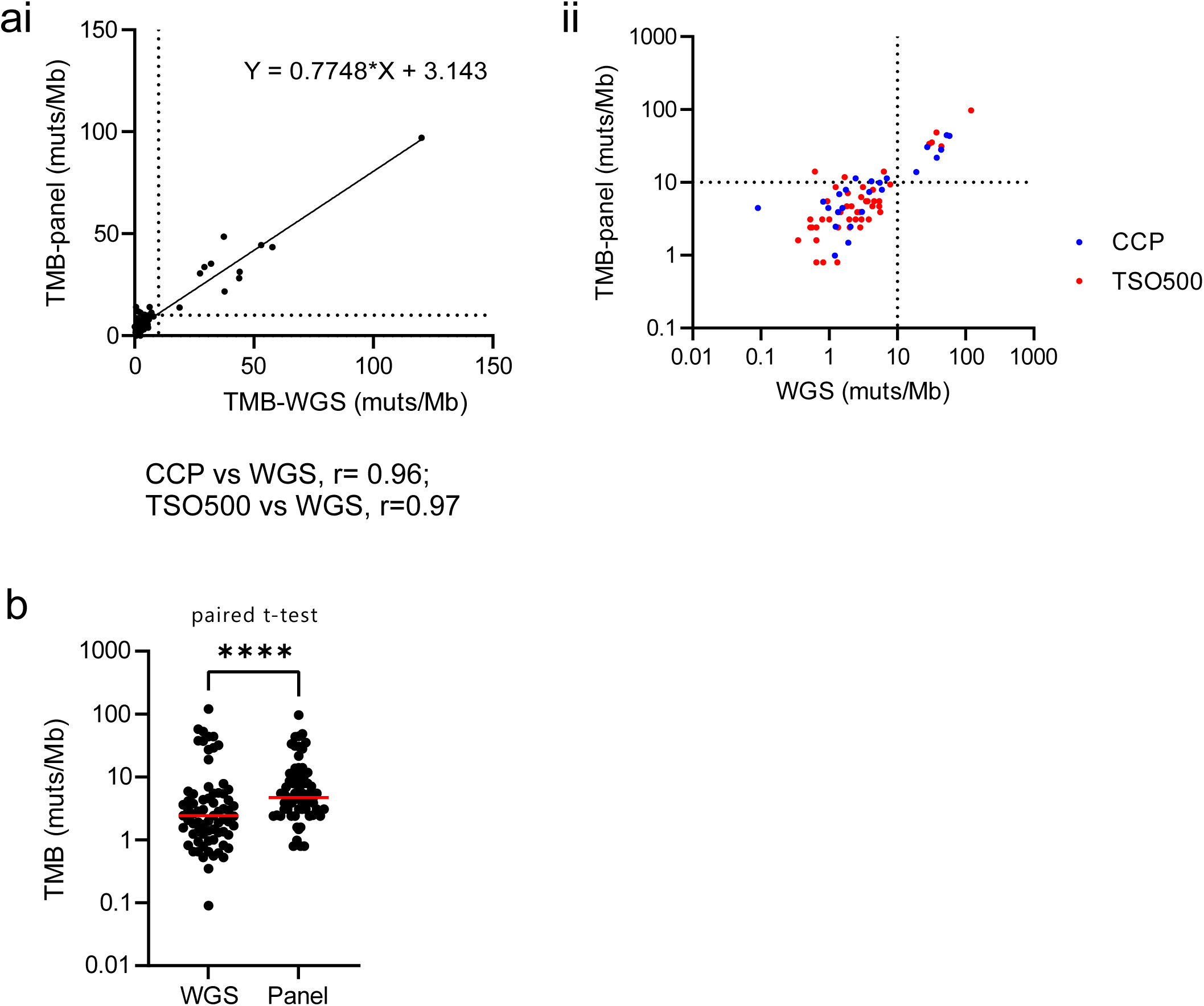
Concordance of TMB estimates in panel and WGS data. **(a)** Correlation of TMB estimates in panel and WGS data across 71 matched cases on **(i)** logarithmic scale and **(ii)** linear scale, showing the Pearson correlation coefficient (r) for each panel-WGS comparison. **(b)** Scatter plot of panel and WGS TMB estimates showing significantly higher TMB estimates in panel than WGS (paired t-test, Wilcoxin rank test, *P*<0.0001).

**Supplementary Figure 2.**
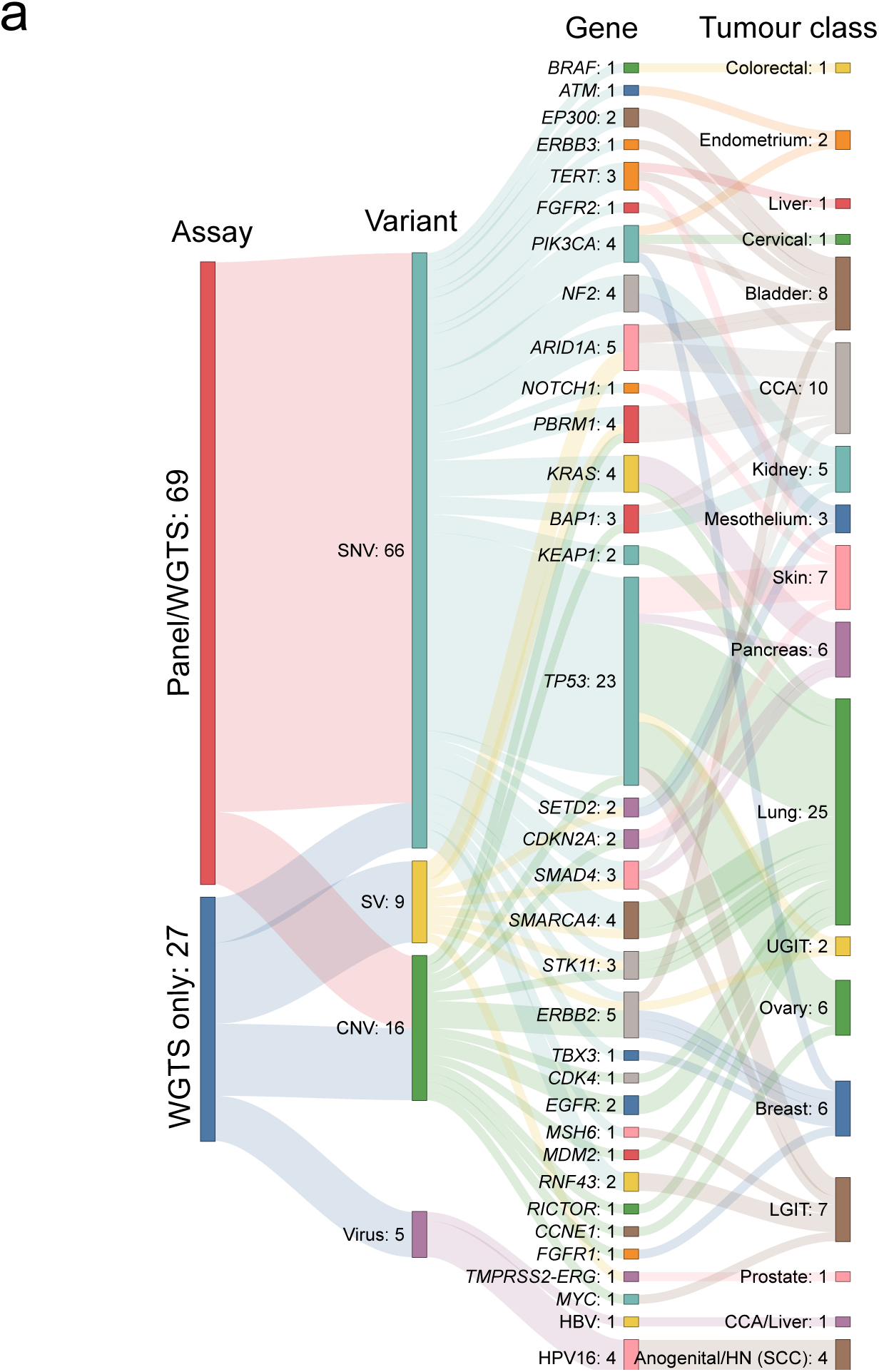
Diagnostic features captured in panel and WGTS data in resolved CUPs. **(a)** Sankey plot of diagnostic features captured in matched panel and WGTS or WGTS only assays, separated by variant type flowing to cancer class.

**Supplementary figure 3.**
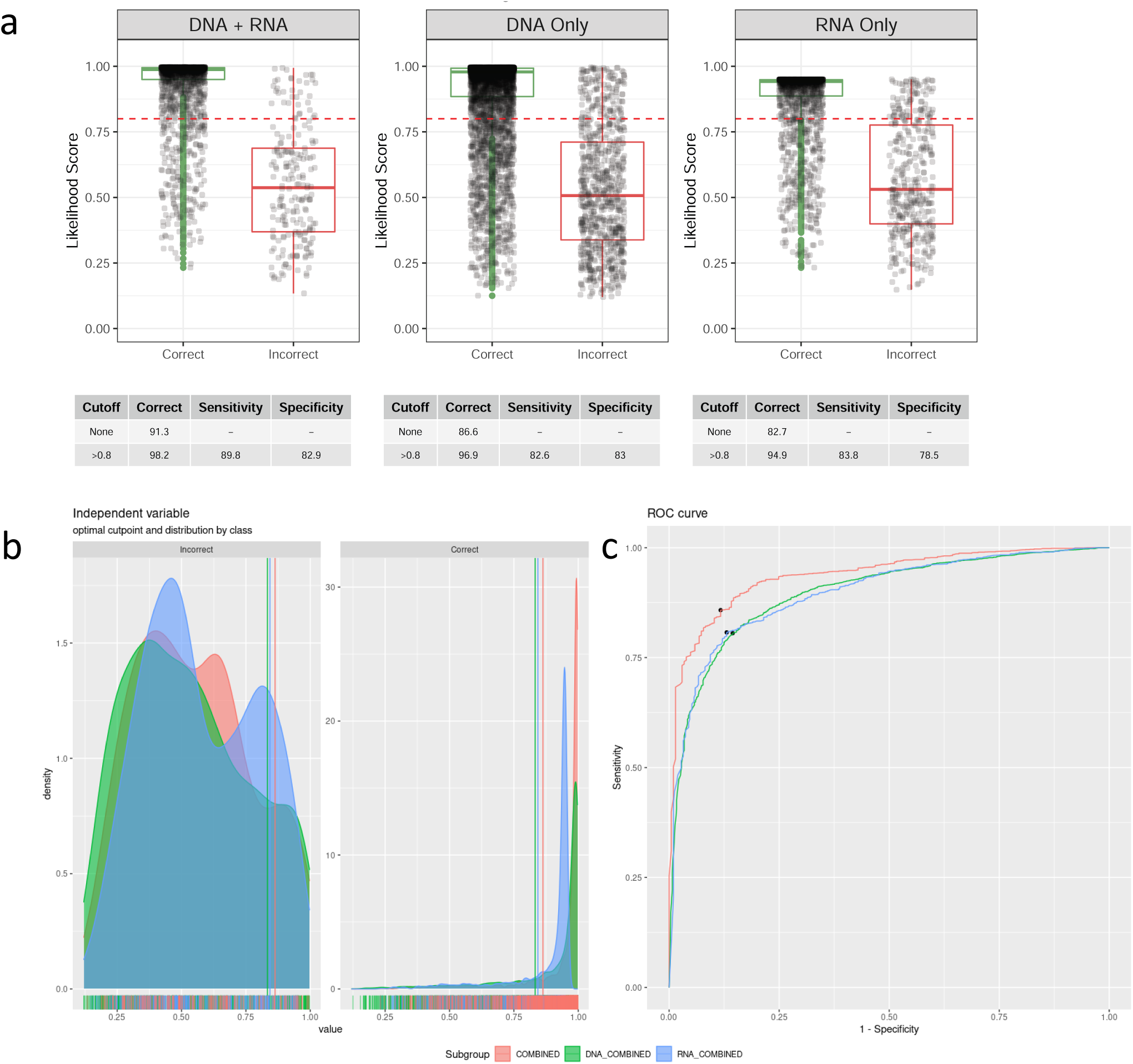
CUPPA DNA, RNA and DNA+RNA classifier validation. **(a)** Box plots grouping correct and incorrect predictions following leave-one-out cross-validation (LOOCV) of CUPPA using a subset of the HMF and PCAWG reference set of WGS (n=6106) and WTS (n=2350) known cancers using DNA + RNA combined, DNA only or RNA only classifiers. Red dotted line indicates the 0.8 likelihood threshold for a high likelihood score. **(b)** Density plot of samples in each classifier showing the optimal cut point and distribution for incorrect predictions (left) and correct predictions (right). Red vertical line indicates the 0.8 likelihood threshold for a high likelihood score. **(c)** Receiver operator curve (ROC) of samples in each classifier showing the optimal cut point of a high likelihood prediction (black dot).

**Supplementary Figure 4.**
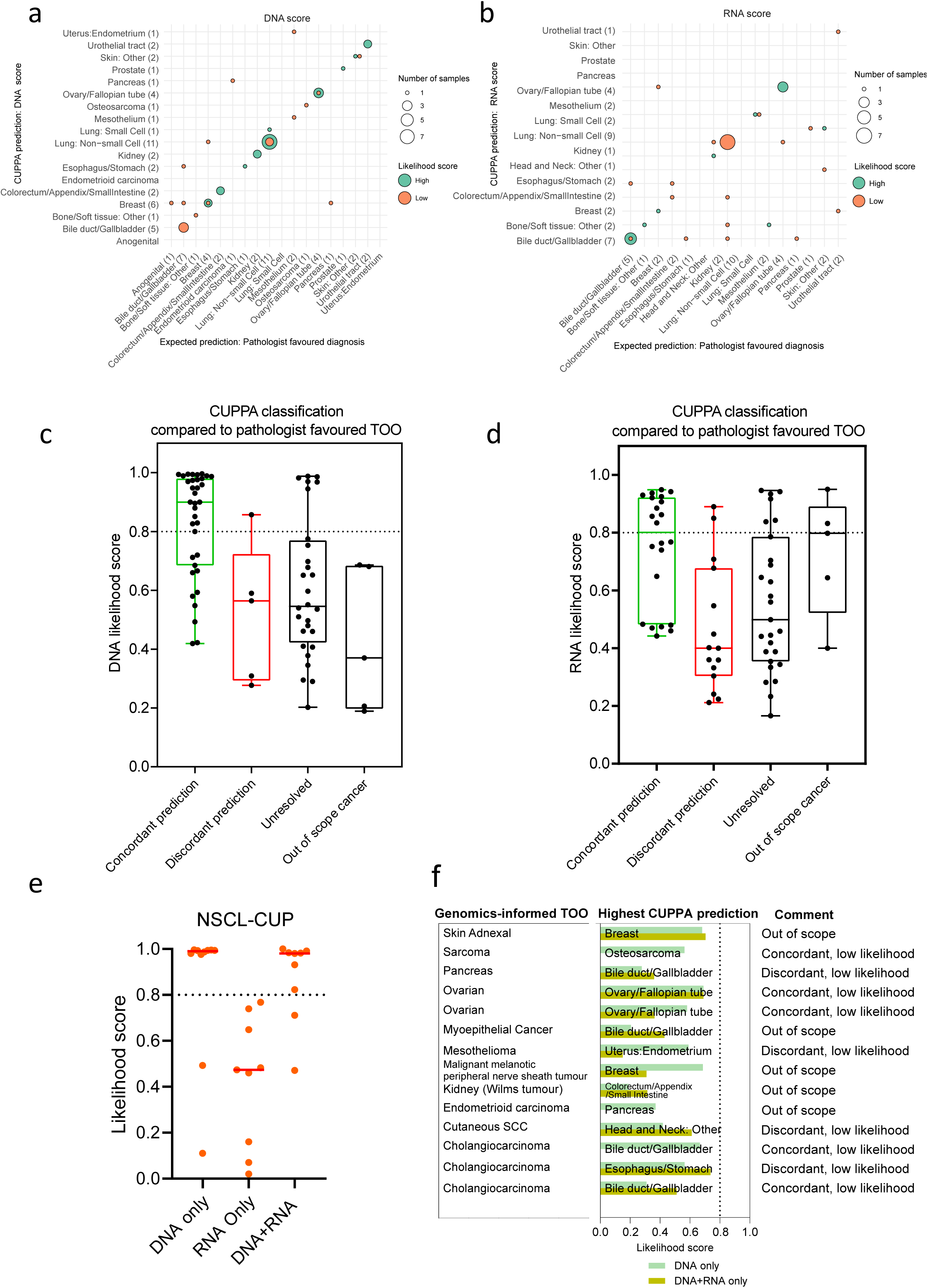
CUPPA DNA only and RNA only predictions in 73 CUP tumours. **(a)** Confusion matrix of DNA only and **(b)** RNA only combined prediction scores against a pathologist’s favoured diagnosis, colour coded by high (≥0.8) or low (<0.8) likelihood. **(c)** Box plot showing combined DNA only and **(d)** RNA only prediction scores for CUPPA across 73 and 69 CUPs, respectively, using classification toward a single site tissue of origin prediction, categorized by concordance with a genomics informed, but CUPPA-blinded, pathology review. Cases with a favoured origin not represented in CUPPA training data are separated as “out of scope cancer”. **(e)** Likelihood scores in resolved non-small cell Lung (NSCL)-CUPs showing DNA only, RNA only and combined DNA+RNA predictions for individual samples. **(f)** Schematic of 14 CUP samples resolved with genomics-informed review that did not achieve a high-likelihood prediction.

**Supplementary Figure 5.**
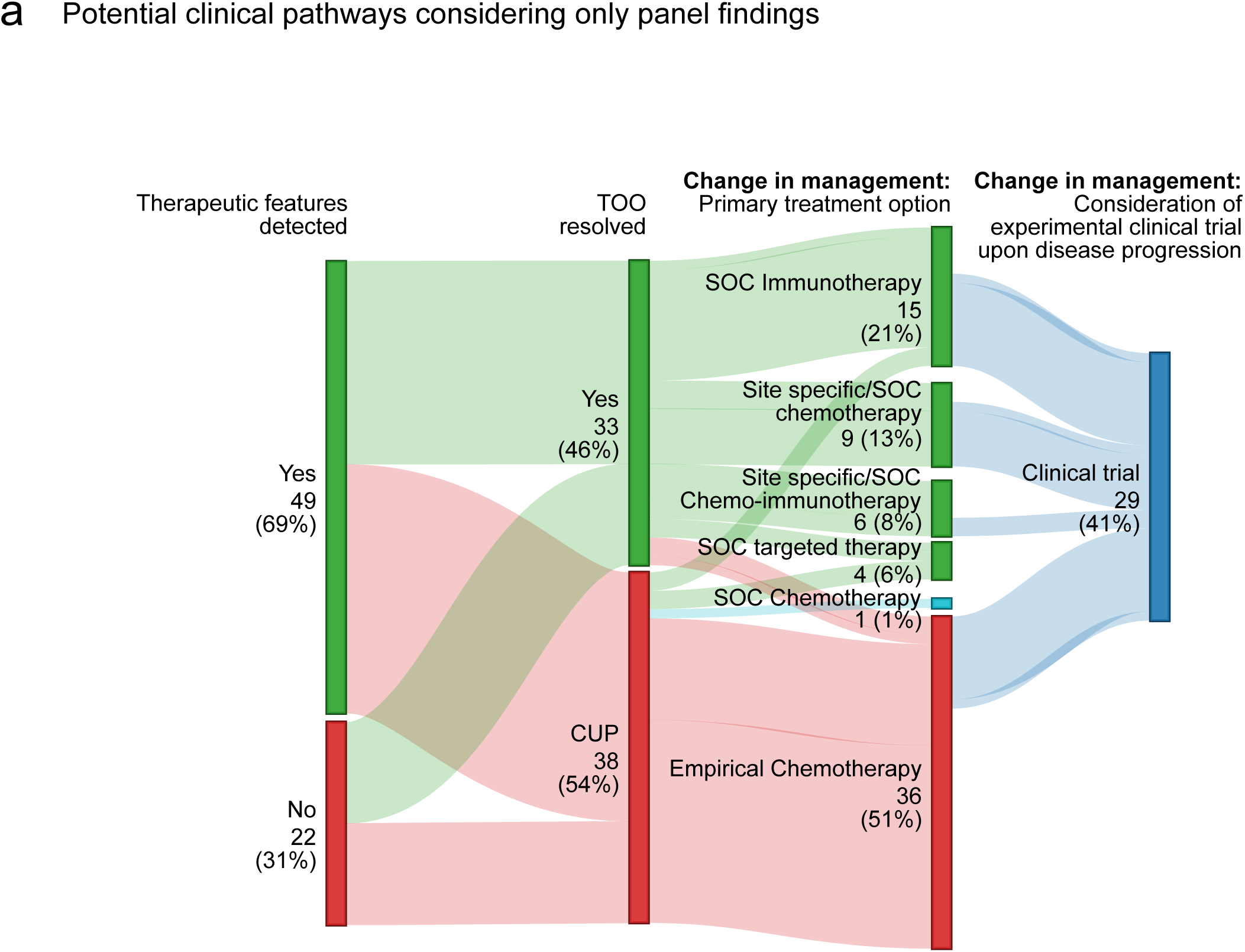
Using panel only derived therapeutic biomarkers and TOO predictions to change clinical management. **(a)** Sankey plot of 71 CUPs showing the number and proportion of cases that had one or more therapeutic biomarkers detected, whether a putative TOO was determined and whether an SOC treatment and/or clinical trial could be considered to gain access to an appropriate treatment.

